# Estimation of Years of Life Lost Due to Premature Mortality from Diabetes Mellitus in Bahia - Brazil

**DOI:** 10.1101/2025.03.24.25324575

**Authors:** Luís Jesuino de Oliveira Andrade, Gabriela Correia Matos de Oliveira, Alcina Maria Vinhaes Bittencourt, João Cláudio Nunes Carneiro Andrade, Janine Lemos de Lima, Luís Matos de Oliveira

## Abstract

**Introduction:** Diabetes mellitus (DM) in Bahia, Brazil, is a critical public health issue, intensified by socioeconomic inequalities. Years of life lost (YLL) highlight healthcare access gaps, impacting vulnerable populations. Enhanced healthcare infrastructure, education, and policy changes are essential to reduce DM morbidity and premature mortality.

**Objective:** To quantify the societal burden of DM in Bahia, Brazil, by estimating YLL due to premature mortality between 2000 and 2023.

**Method:** This study uses a quantitative epidemiological approach to estimate YLL due to DM in Bahia, Brazil (2000–2023). Mortality data from the Mortality Information System (SIM) and demographic projections from IBGE were analyzed. YLL was calculated by multiplying deaths by remaining life expectancy (using GBD 2019 reference tables), stratified by age and sex. Statistical analysis employed PSPP, Excel, and Python, with results presented as rates per 100,000 population. Socioeconomic and healthcare factors were contextualized using government reports and academic literature.

**Results:** The study analyzed YLL due to DM in Bahia (2000–2023), revealing an upward trend, peaking in 2020 (422.6/100,000), likely exacerbated by COVID-19. Males showed higher premature mortality than females (34.9 vs. 29.8/100,000 in 2020). Economic losses reached ~41 million BRL (2020–2022). Rising YLL reflects growing DM burden, with socioeconomic impacts including lost productivity and household instability. Gender disparities suggest differences in healthcare access, biological susceptibility, or lifestyle factors, necessitating targeted interventions.

**Conclusion:** Bahia faces rising diabetes-related premature deaths, worsening post-2015 and spiking during COVID-19. Men show higher mortality, with severe socioeconomic impacts, requiring urgent public health and economic strategies.

## INTRODUCTION

Diabetes mellitus (DM) is an important public health challenge globally, with its burden disproportionately affecting low- and middle-income regions such as Bahia, Brazil.^1^ As a metabolic disorder characterized by chronic hyperglycemia, DM contributes significantly to premature mortality through complications like cardiovascular disease, renal failure, and neuropathy. Between 2000 and 2024, Bahia a state marked by socioeconomic disparities and uneven healthcare access has experienced rising DM prevalence, mirroring national trends but compounded by regional inequities.^2^ Quantifying the societal impact of DM-related deaths requires robust epidemiological metrics, such as Years of Life Lost (YLL), which estimate the gap between observed age at death and life expectancy.

YLL serves as an important tool for contextualizing premature mortality, emphasizing the societal cost of diseases like DM beyond crude mortality rates.^3^ Recent studies in Brazil highlight YLL’s utility in uncovering hidden burdens in populations with fragmented health data, particularly in the Northeast region, where Bahia is situated.^4^ For instance, was demonstrated that YLL calculations in similar settings reveal up to 40% higher disease impacts than traditional mortality analyses, underscoring the need for granular, region-specific assessments.^5^ This approach is especially pertinent in Bahia, where systemic gaps in chronic disease management exacerbate preventable DM complications.

Socioeconomic determinants, including income inequality and limited primary care coverage, disproportionately affect DM outcomes in Bahia. A cohort study linked low socioeconomic status to a 2.3-fold increase in DM-related mortality among Bahian adults under 60, highlighting premature death as a marker of healthcare inequity.^6^ Furthermore, urbanization and shifts toward sedentary lifestyles have amplified DM incidence, yet public health responses remain fragmented. Chaves-Fonseca RM, et al.^7^ noted that less than 60% of Bahia’s municipalities met national targets for diabetes care infrastructure, perpetuating regional disparities in mortality.

Temporal trends in DM mortality further complicate Bahia’s landscape. While national data show a 22% decline in DM-related YLL from 2010 to 2019, Bahia’s reduction lagged at 12%, attributed to slower implementation of preventive strategies.^8^ Emerging evidence also suggests that the COVID-19 pandemic disrupted routine DM management, potentially exacerbating premature mortality post-2020.^9^

This study’s findings aim to inform targeted interventions by delineating the magnitude and drivers of DM-related YLL in Bahia. By integrating mortality data with demographic projections, the analysis will address gaps identified in the Global Burden of Disease framework, which often overlooks subnational heterogeneity.^10^ For Bahia home to 14.850.513 people, such granularity is critical for aligning resource allocation with the UN’s Sustainable Development Goals, particularly Target 3.4, which prioritizes reducing premature mortality from noncommunicable diseases.^11^

This study seeks to quantify the societal burden of DM in Bahia, Brazil, by estimating YLL due to premature mortality between 2000 and 2023. By integrating mortality data with demographic and epidemiologic projections, the analysis aims to elucidate the magnitude of preventable DM-related deaths, contextualizing disparities driven by socioeconomic inequities, fragmented healthcare access, and evolving public health challenges.

## METHODOLOGY

This study will employ a quantitative, epidemiological approach to estimate YLL attributable to DM in Bahia, Brazil, between 2000 and 2023. The methodology will involve a multi-stage process integrating mortality data, demographic projections, and relevant epidemiological parameters.

### Data Sources

Mortality data for DM spanning the period from 2000 to 2023 were obtained through the Mortality Information System (SIM), developed by the Brazilian Ministry of Health in 1975. This system resulted from the unification of over forty different instrument models used throughout the years to collect mortality data in the country (http://tabnet.datasus.gov.br/cgi/deftohtm.exe?sim/cnv/obt10uf.def). These data were stratified by age, sex, and underlying cause of death, specifically focusing on deaths where DM is listed as the primary or contributing cause. Demographic data, including population counts and age-sex distributions for Bahia, were obtained from the Brazilian Institute of Geography and Statistics (IBGE). These data were used to project the population at risk of DM-related mortality throughout the study period. Epidemiological parameters, such as DM prevalence rates, were drawn from national health surveys and published literature specific to the Bahian population.

### YLL Calculation

YLL was calculated using the standard methodology, which involves subtracting the age at death from a standard life expectancy:

Basic Formula: YLL = Σ (Number of deaths in each age group) × (Life expectancy in that age group)

The choice of standard life expectancy was carefully considered, potentially utilizing the World Health Organization (WHO) standard or a Brazilian national life expectancy table, ensuring consistency and comparability with other studies. YLL was calculated for each death attributed to DM and then aggregated to provide an overall estimate of DM-related YLL for Bahia. The standard life expectancy was projected based on the mean age at death, calculated with a 95% confidence interval. For this purpose, the global life table from the GBD (Global Burden of Disease Study) 2019 study was used as a reference to ensure comparability with existing studies.

### Statistical Analysis

YLL due to premature mortality, as defined by the GBD study methodology, addresses the limitations of arbitrary age thresholds by quantifying lost time rather than raw death counts, using an individual’s maximum potential lifespan at each age as the foundational metric.^12^ YLL is calculated by multiplying the number of deaths by the remaining life expectancy at the age of death. For DM, sex, and age group stratification, YLL estimates were derived as follows:

The core formula for YLL for a given cause *c*, sex *s*, age *a*, and year *t* is:

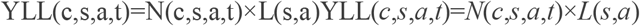

where:

- N(c,s,a,t)*N*(*c,s,a,t*) = number of deaths due to cause *c* for sex *s* and age *a* in year *t*;
- L(s,a)*L*(*s,a*) = standard loss function specifying years of life lost per death at age *a* for sex *s*.

As described by Murray et al.,^13^ a discount rate may be applied to YLL to prioritize present health benefits over future ones, effectively reducing the value of each year of life by a fixed annual percentage. For this study, a 3% discount rate was not adopted.

Regarding age weighting and time discounting, the 1990 GBD study by the WHO calculated Disability-Adjusted Life Years (DALY) using a 3% discount rate for future healthy life years lost, alongside an alternative 0% rate. Proponents of discounting argued its necessity to avoid decision-making paradoxes in cost-effectiveness analyses.^14^ Critics, however, contended that there is no ethical justification for devaluing future health outcomes,^15^ a stance reinforced by expert consultations for the 2010 GBD study, which discouraged discounting.^16^

Data processing and variable management were performed using PSPP Statistics (open-source, public domain software). YLL calculations were executed in Excel (Office 2019 suite), the WHO deterministic DALY calculation template (available: http://www.who.int/healthinfo/bodreferencedalycalculationtemplate.xls), and Python-Fiddle (https://python-fiddle.com/?checkpoint=1742657084). Results are presented as rates per 100,000 population, stratified by sex and age group.

Productivity loss in this study is analyzed through the human capital theory framework, which views workers as economic investments generating returns. Productivity loss refers to diminished capacity of individuals or groups to engage in economically productive activities due to health impairments or adverse conditions.

The economic cost of premature mortality due to productivity loss in Bahia was estimated using the following indicators: Expected annual income (RE); YLL; A 5% discount rate.

The formula for economic loss due to reduced productivity is:

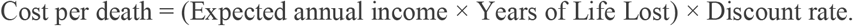

### Socioeconomic and Healthcare Contextualization

To contextualize the observed YLL trends, data on socioeconomic indicators (e.g., income inequality, poverty rates), healthcare access (e.g., primary care coverage, availability of diabetes care facilities), and public health initiatives related to DM prevention and management in Bahia were gathered from relevant sources, such as government reports, academic publications, and publicly available databases. Qualitative contextualization was also be considered, drawing upon existing literature to discuss the complex interplay of social determinants, healthcare system factors, and public health policies influencing DM-related mortality in Bahia.

### Ethical Considerations

This study utilized publicly available, anonymized data, and therefore, individual patient consent was not be required. So, according to Article 1, Sole Paragraph, of Resolution No. 510/2016, the following will not be registered or evaluated by the Research Ethics Committee - Brazil: Research using databases in which the information is aggregated, with no possibility of individual identification.

## RESULTS

### Demographic data of population of Bahia

The demographic data for the population of Bahia were sourced from the Health Information System of the Bahia State Health Department, with population estimates compiled from the IBGE for the resident population in the region for the year 2020, as outlined in Table 1.

**Table 1.**
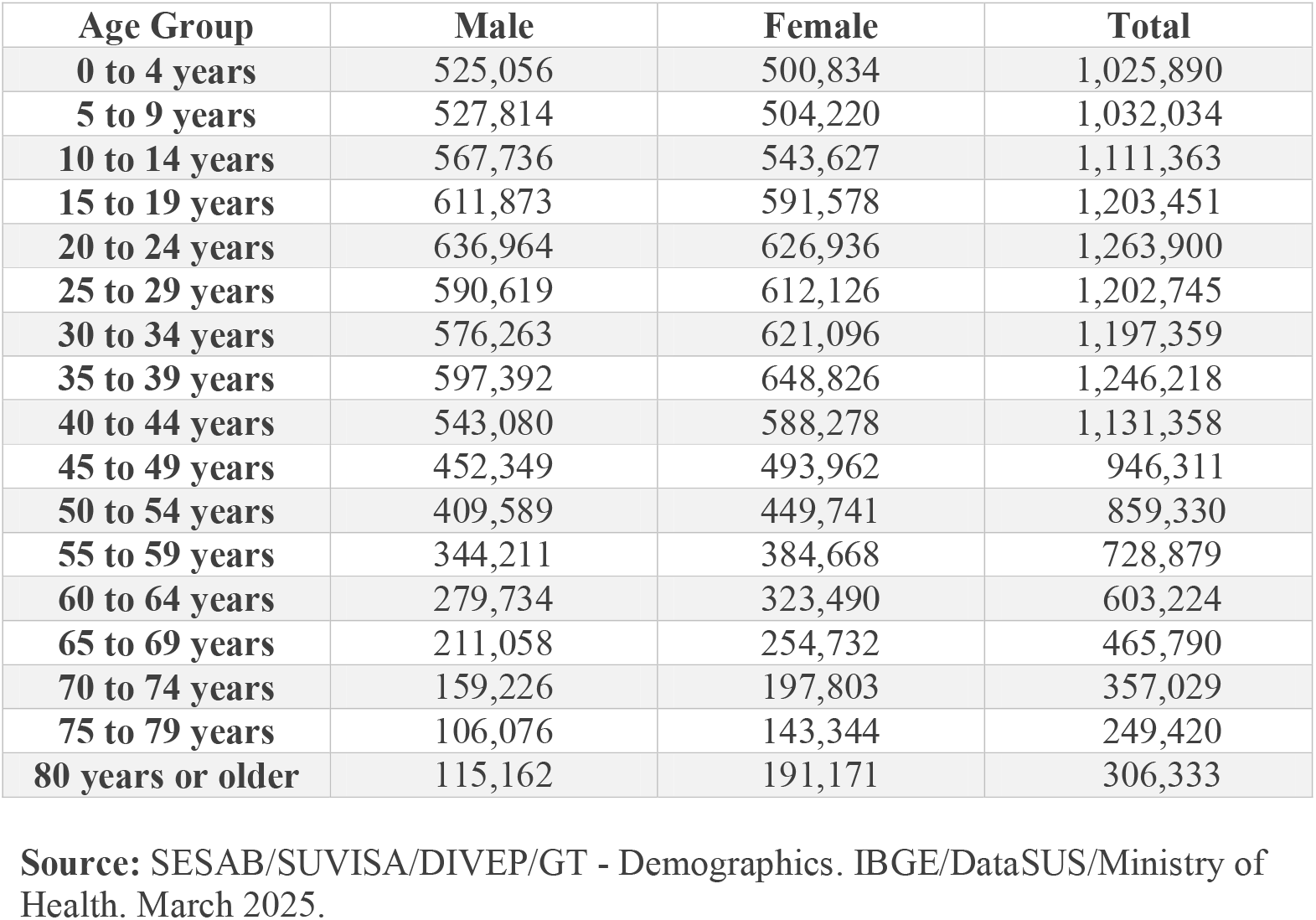
Estimated resident population, stratified by sex and age group – Bahia, 2020 to 2022. **Source:** SESAB/SUVISA/DIVEP/GT - Demographics. IBGE/DataSUS/Ministry of Health. March 2025.

### Standard life expectancy

At present, the most recent iteration of the GBD study accessible for investigative purposes is the GBD 2019. This comprehensive research endeavor is conducted by the Institute for Health Metrics and Evaluation (IHME) at the University of Washington, with data undergoing periodic updates to reflect evolving health landscapes. The GBD 2019 provides extensive evaluations concerning mortality, morbidity, risk determinants, and life expectancy across more than 200 nations and territories, encapsulating the period from 1990 to 2019.

Key Attributes of the GBD 2019: 1) Broad Data Spectrum: It encompasses evaluations for over 350 distinct diseases and injuries, supplemented by 84 identified risk factors; 2) Demographic Segmentation: The dataset is organized by age brackets, gender, and geographical location to facilitate granular analysis; 3) Health Metric Inclusion: It integrates DALY, YLL, and Years Lived with Disability (YLD) as core health metrics; 4) Risk Factor Analysis: It scrutinizes the influence of variables such as obesity, tobacco consumption, and atmospheric pollution on the global disease burden.

Access to the data was made through the IHME website: https://www.healthdata.org/gbd/2019.

The standard life expectancy was projected based on the average age at death, calculated with a 95% confidence interval. For this purpose, the Global Life Table from the GBD 2019 study was used as a reference to ensure comparability with other previously conducted studies (Table 2 and 3).

**Table 2.** Global Life Expectancy, 2019.

| AGE GROUP | 2019 |  |
| --- | --- | --- |
|  | Male | Female |
| Under 1 year | 70.85 | 75.87 |
| 1-4 years | 72.03 | 76.88 |
| 5-9 years | 68.70 | 73.59 |
| 10-14 years | 63.96 | 68.86 |
| 15-19 years | 59.16 | 64.04 |
| 20-24 years | 54.43 | 59.23 |
| 25-29 years | 49.79 | 54.47 |
| 30-34 years | 45.17 | 49.72 |
| 35-39 years | 40.59 | 45.00 |
| 40-44 years | 36.10 | 40.34 |
| **45-49 years** | 31.71 | 35.75 |
| **50-54 years** | 27.43 | 31.23 |
| **55-59 years** | 23.34 | 26.87 |
| **60-64 years** | 19.52 | 22.67 |
| **65-69 years** | 15.99 | 18.71 |
| **70-74 years** | 12.74 | 14.99 |
| **75-79 years** | 9.83 | 11.62 |
| **80-84 years** | 7.26 | 8.62 |
| **85+ Years** | 5.13 | 6.15 |

**Source:** GBD, 2019.

**Table 3.**
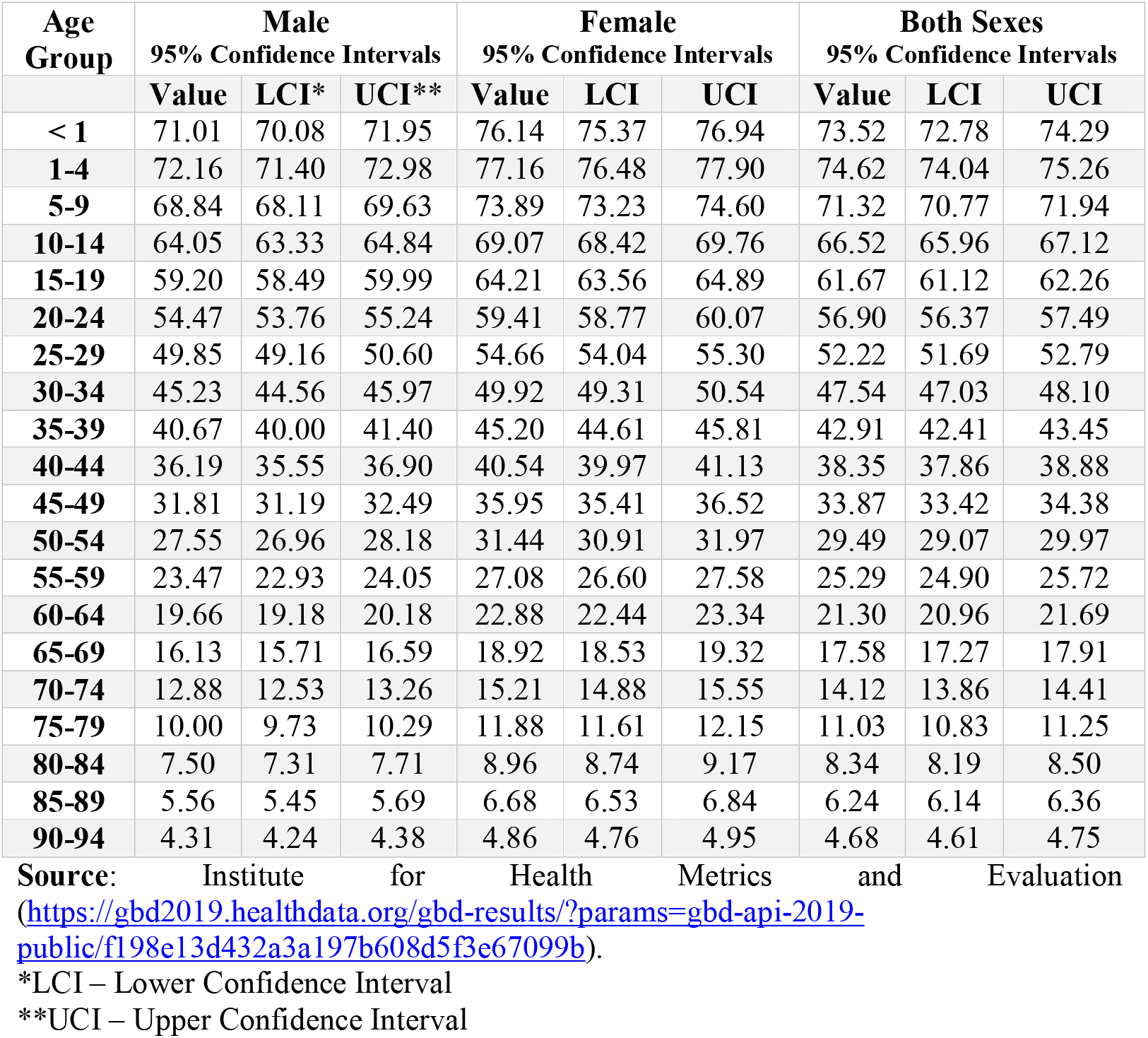
Age-Specific Data by Sex with 95% Confidence Intervals. **Source**: Institute for Health Metrics and Evaluation (https://gbd2019.healthdata.org/gbd-results/?params=gbd-api-2019-public/f198e13d432a3a197b608d5f3e67099b).

| **Age Group** | **Male** |  |  | **Female** |  |  | **Both Sexes** |  |  |
| --- | --- | --- | --- | --- | --- | --- | --- | --- | --- |
|  | **95% Confidence Intervals** |  |  | **95% Confidence Intervals** |  |  | **95% Confidence Intervals** |  |  |
|  | **Value** | **LCI\*** | **UCI\*\*** | **Value** | **LCI** | **UCI** | **Value** | **LCI** | **UCI** |
| **< 1** | 71.01 | 70.08 | 71.95 | 76.14 | 75.37 | 76.94 | 73.52 | 72.78 | 74.29 |
| **1-4** | 72.16 | 71.40 | 72.98 | 77.16 | 76.48 | 77.90 | 74.62 | 74.04 | 75.26 |
| **5-9** | 68.84 | 68.11 | 69.63 | 73.89 | 73.23 | 74.60 | 71.32 | 70.77 | 71.94 |
| **10-14** | 64.05 | 63.33 | 64.84 | 69.07 | 68.42 | 69.76 | 66.52 | 65.96 | 67.12 |
| **15-19** | 59.20 | 58.49 | 59.99 | 64.21 | 63.56 | 64.89 | 61.67 | 61.12 | 62.26 |
| **20-24** | 54.47 | 53.76 | 55.24 | 59.41 | 58.77 | 60.07 | 56.90 | 56.37 | 57.49 |
| **25-29** | 49.85 | 49.16 | 50.60 | 54.66 | 54.04 | 55.30 | 52.22 | 51.69 | 52.79 |
| **30-34** | 45.23 | 44.56 | 45.97 | 49.92 | 49.31 | 50.54 | 47.54 | 47.03 | 48.10 |
| **35-39** | 40.67 | 40.00 | 41.40 | 45.20 | 44.61 | 45.81 | 42.91 | 42.41 | 43.45 |
| **40-44** | 36.19 | 35.55 | 36.90 | 40.54 | 39.97 | 41.13 | 38.35 | 37.86 | 38.88 |
| **45-49** | 31.81 | 31.19 | 32.49 | 35.95 | 35.41 | 36.52 | 33.87 | 33.42 | 34.38 |
| **50-54** | 27.55 | 26.96 | 28.18 | 31.44 | 30.91 | 31.97 | 29.49 | 29.07 | 29.97 |
| **55-59** | 23.47 | 22.93 | 24.05 | 27.08 | 26.60 | 27.58 | 25.29 | 24.90 | 25.72 |
| **60-64** | 19.66 | 19.18 | 20.18 | 22.88 | 22.44 | 23.34 | 21.30 | 20.96 | 21.69 |
| **65-69** | 16.13 | 15.71 | 16.59 | 18.92 | 18.53 | 19.32 | 17.58 | 17.27 | 17.91 |
| **70-74** | 12.88 | 12.53 | 13.26 | 15.21 | 14.88 | 15.55 | 14.12 | 13.86 | 14.41 |
| **75-79** | 10.00 | 9.73 | 10.29 | 11.88 | 11.61 | 12.15 | 11.03 | 10.83 | 11.25 |
| **80-84** | 7.50 | 7.31 | 7.71 | 8.96 | 8.74 | 9.17 | 8.34 | 8.19 | 8.50 |
| **85-89** | 5.56 | 5.45 | 5.69 | 6.68 | 6.53 | 6.84 | 6.24 | 6.14 | 6.36 |
| **90-94** | 4.31 | 4.24 | 4.38 | 4.86 | 4.76 | 4.95 | 4.68 | 4.61 | 4.75 |

\*LCI – Lower Confidence Interval
\*\*UCI – Upper Confidence Interval

### YLL Attributable to Premature Mortality from DM in Bahia, Brazil

The analyses revealed the following YLL attributable to premature mortality from DM in Bahia: 2000: 151.7/100,000 inhabitants; 2001: 178.9/100,000 inhabitants; 2002: 188.0/100,000 inhabitants; 2003: 190.9/100,000 inhabitants; 2004: 196.6/100,000 inhabitants; 2005: 189.2/100,000 inhabitants; 2006: 234.9/100,000 inhabitants; 2007: 247.3/100,000 inhabitants; 2008: 262.0/100,000 inhabitants; 2009: 272.3/100,000 inhabitants; 2010: 284.6/100,000 inhabitants; 2011: 307.4/100,000 inhabitants; 2012: 309.4/100,000 inhabitants; 2013: 309.6/100,000 inhabitants; 2014: 312.0/100,000 inhabitants; 2015: 330.0/100,000 inhabitants; 2016: 314.3/100,000 inhabitants; 2017: 347.8/100,000 inhabitants; 2018: 341.0/100,000 inhabitants; 2019: 341.0/100,000 inhabitants; 2020: 422.6/100,000 inhabitants; 2021: 419,8/100,000 inhabitants; 2022: 437,9/100,000 inhabitants; and 2023: 431,2/100,000 inhabitants (Graph 1).

**Graph 1.**
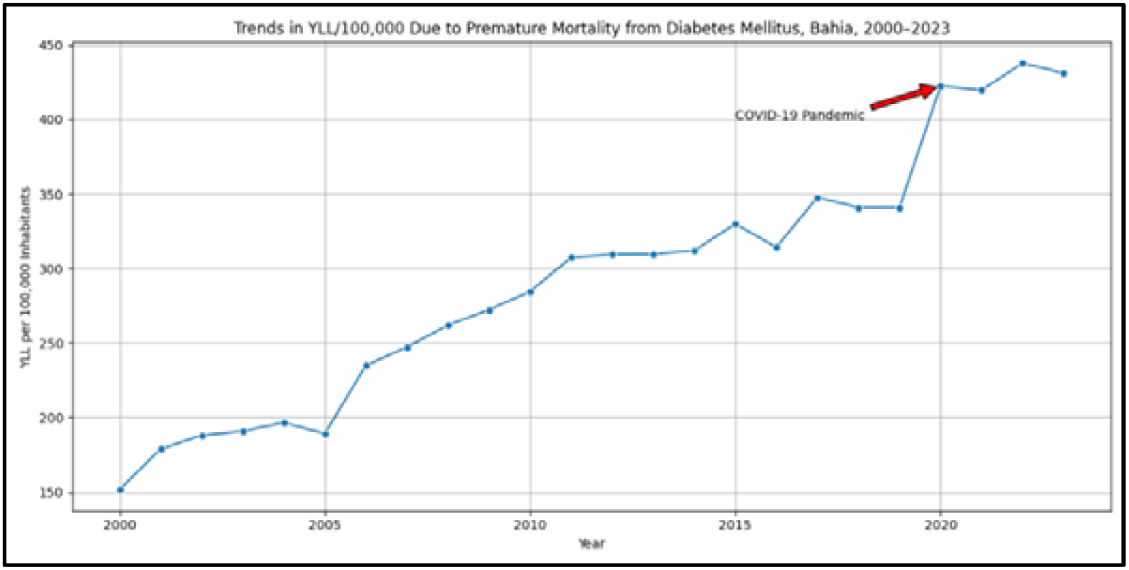
Attributable to Premature Mortality from DM in Bahia **Source**: Study result

The increasing trend in YLL/100,000 indicates a growing burden of premature mortality due to DM in Bahia over the past two decades, with notable peaks in 2020. A sharp rise is observed between 2005 and 2007, followed by a plateau from 2012 to 2019. The spike in 2020 suggests that external factors, such as the COVID-19 pandemic, may have contributed to increased diabetes-related deaths.

### Interpretive insights

#### Accelerated growth

Non-linear trend shows increasing slope after 2015 (^β^2015-2023 > ^β^2000-2015).

#### Pandemic amplification

24.2% YLL increase from 2019 to 2020 (Δ=81.6/100kΔ=81.6/100k).

#### Sustained burden

Post-pandemic rates remain 26.4% above pre-2020 baseline.

### Gender Disparity in YLL Due to Premature Diabetes Mellitus Mortality

The age-standardized premature mortality rates, per 100,000 inhabitants, due to DM among individuals aged 30–69 years in Bahia, stratified by gender, reveal significant trends and disparities over the period from 2000 to 2023. Among females, rates fluctuated moderately, peaking at 31.4 in 2009 and showing a slight increase to 29.8 in 2020, likely influenced by the COVID-19 pandemic. In contrast, males exhibited higher and more pronounced rates, with a sharp peak of 34.9 in 2020, reflecting a greater vulnerability to diabetes-related complications during health crises. While both genders experienced a decline in mortality rates after 2009, males consistently maintained higher rates, with a notable upward trend from 2014 onward, reaching 32.4 in 2015 and 34.5 in 2023. Females, on the other hand, demonstrated relative stability, with rates hovering around 28–29 from 2016 to 2023. The data suggests that males face a disproportionately higher burden of premature mortality due to Diabetes Mellitus, potentially due to differences in healthcare access, lifestyle factors, or biological susceptibility, as detailed in Graph 2 and Graph 3.

**Graph 3.**
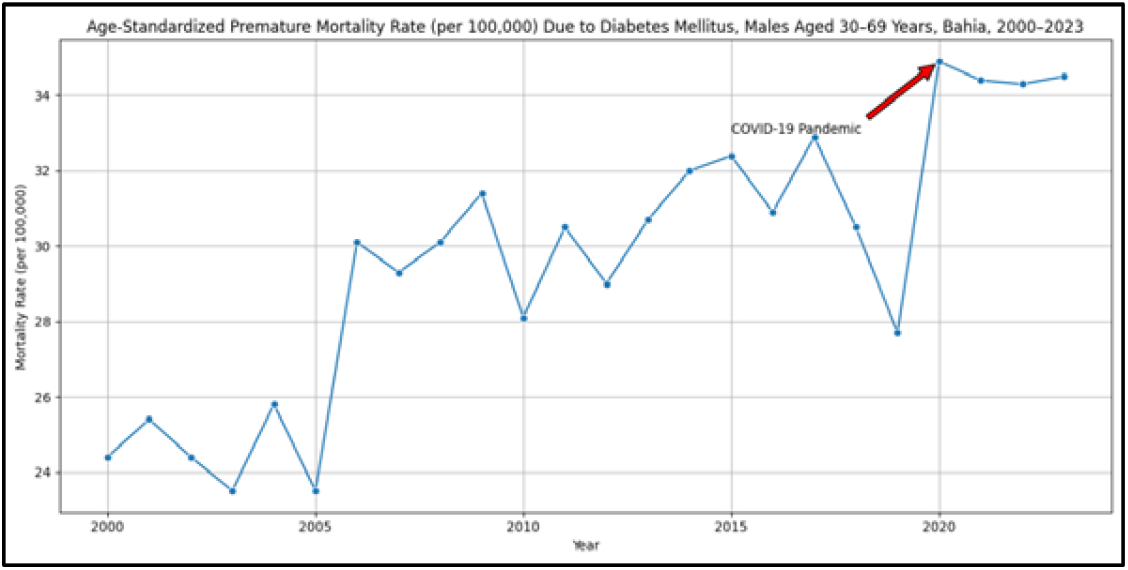
YLL Due to Premature Death, DM, Bahia, Brazil 2000-2023, Male. **Source**: Study result

**Graph 2.**
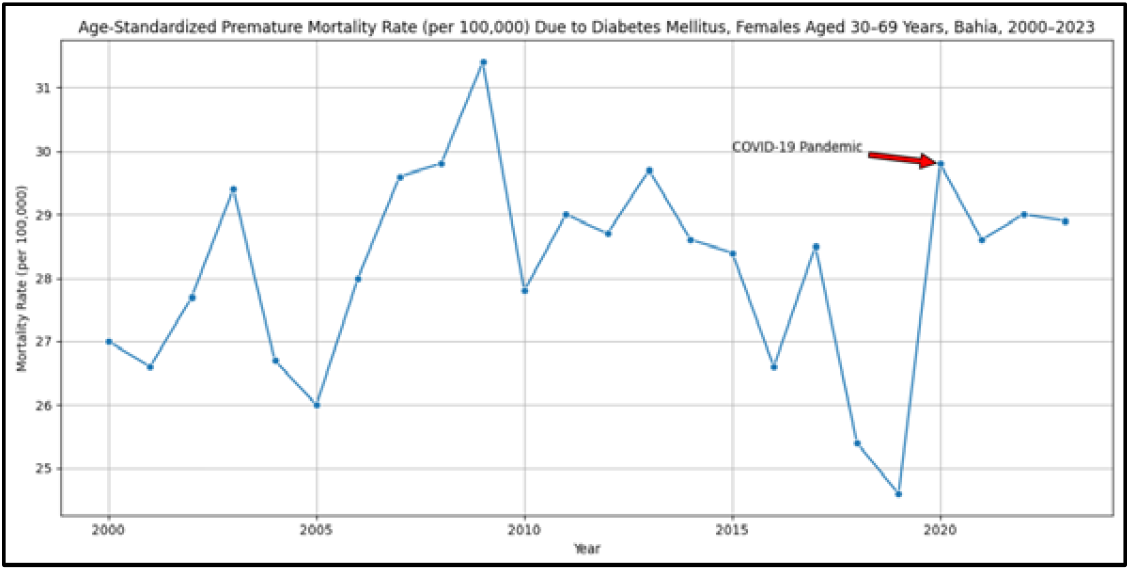
YLL Due to Premature Death, DM, Bahia, Brazil 2000-2023, Female. **Source**: Study result

### Socioeconomic Implications of Premature Diabetes Mellitus Mortality in Bahia

In 2020, the per capita income in Bahia was R$ 1,568, and in 2022, it was R$ 1,613.^17^ Consequently, the economic loss attributable to premature mortality due to diabetes mellitus was approximately 41 million Brazilian Reais.

The escalating premature mortality due to DM in Bahia, evidenced by the increasing trend in YLL per 100,000 inhabitants, carries profound socioeconomic implications for the state. Over the past 20 years, the surge in YLL, particularly the sharp increase observed between 2005 and 2007 and the peak in 2020, underscores the growing impact of DM on the working-age population. The peak in 2020, likely exacerbated by the COVID-19 pandemic, highlights how health crises can amplify the vulnerability of individuals with chronic conditions such as diabetes. Premature deaths among individuals aged 30 to 69 not only represent a tragic loss of lives but also deprive families of their primary providers, disrupt household stability, and place additional financial strain on surviving members. This loss of productivity and increased reliance on social support systems further exacerbate economic inequalities, particularly in low- and middle-income regions like Bahia (Graph 4).

**Graph 4.**
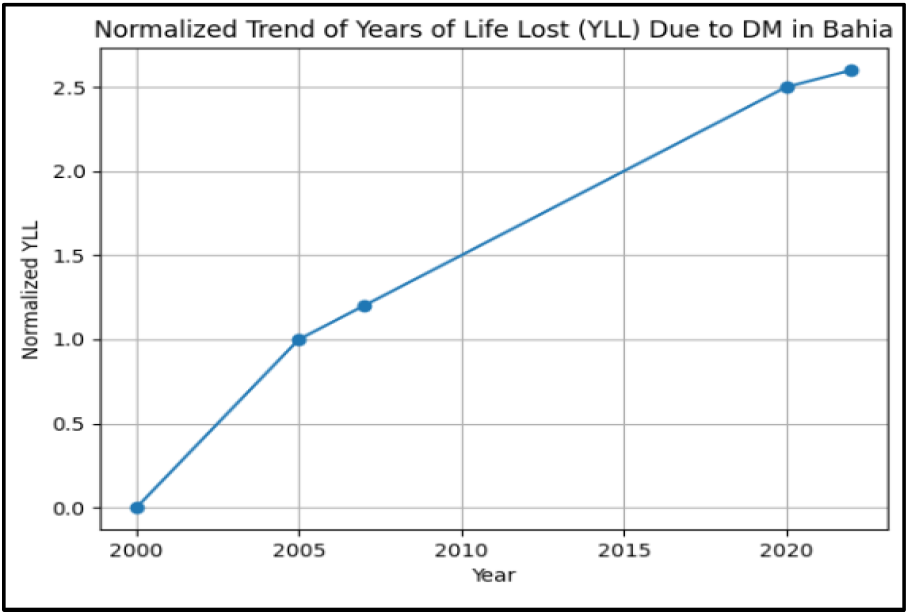
Trend YLL due to DM in Bahia. **Source**: Study result

The gender disparities in premature mortality rates reveal a disproportionate impact between men and women, with rates peaking at 34.9 in 2020 and remaining elevated through 2023 (Graph 5).

**Graph 5.**
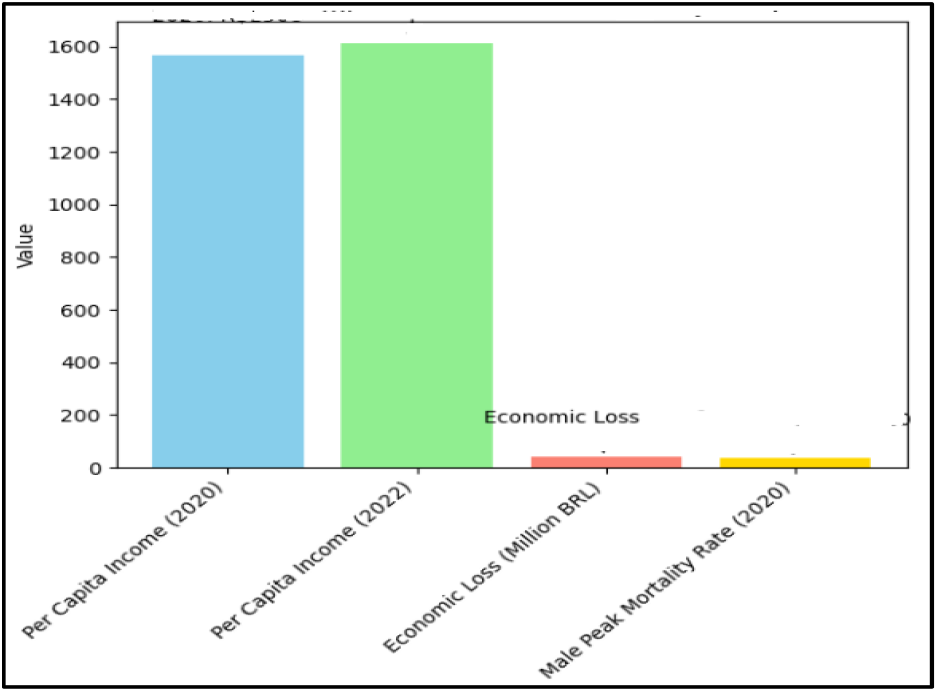
The gender disparities in premature mortality rates. **Source**: Study result

This trend suggests that men may face greater challenges in accessing healthcare, adopting preventive measures, or managing DM-related complications, potentially due to social norms, occupational hazards, or biological factors. In contrast, women, although also affected, exhibit more stable mortality rates, indicating potential resilience or better engagement with health services. Addressing the socioeconomic impact of premature DM mortality requires a comprehensive approach that integrates public health strategies, economic support, and gender-sensitive policies to attenuate the long-term consequences on families, communities, and the broader economy.

## DISCUSSION

Our time-series analysis of premature DM mortality in Bahia, Brazil, reveals a concerning trend of increasing YLL over the of more than two decades. This pattern underscores the significant impact of DM on public health, particularly in terms of premature mortality. The escalating YLL rates suggest that DM continues to pose a substantial problem for the healthcare system and the community, highlighting the need for targeted interventions and enhanced care strategies. Demographic characteristics such as age, sex, and socioeconomic factors play a significant role in understanding the dynamics of DM-related mortality. The observed trajectory of the study’s results, marked by a steady climb and punctuated by mortality spikes, demands a deeper exploration of the underlying factors driving this public health issue.

Estimates of YLL represent a critical metric in public health, quantifying the impact of premature mortality by calculating the difference between the age at death and a predetermined standard life expectancy.^18^ This index surpasses crude mortality rates, offering nuanced insights into the burden of diseases or injuries, particularly emphasizing deaths occurring at younger ages.^19^ Variations in standard life expectancy across studies may affect comparability, necessitating methodological transparency and context-specific interpretations. Studies suggest that YLL varies significantly across demographic data, highlighting disparities in healthcare access and socioeconomic factors.^20^ YLL has been deemed indispensable for prioritizing public health interventions and resource allocation.^21^ This study evaluated the YLL attributable to premature mortality from 2000 to 2023 due to DM, aiming to assess the impact of DM on population health. The evaluation focused on quantifying the YLL and its implications for public health policy, which can provide important insights into the disease burden and thereby lead to the development of strategies for targeted interventions according to public health services.

The demography of Bahia, a state with vast socioeconomic and cultural diversity, presents a complex population profile. With an estimated population exceeding 14 million inhabitants, Bahia is the fourth most populous state in Brazil, characterized by an uneven distribution, featuring large urban concentrations and extensive rural areas.^22^ The ongoing demographic transition, marked by population aging and declining birth rates, poses significant challenges for public health and social security policies.^23^ Furthermore, the high prevalence of non-communicable chronic diseases, such as DM and hypertension, necessitates targeted interventions for health promotion and disease prevention.^24^ In our study, demographic data for the Bahian population were sourced from the Health Information System of the Bahia State Health Department, with population estimates systematically compiled by the IBGE. This approach ensured the utilization of authoritative and comprehensive datasets, facilitating robust demographic analysis and enhancing the reliability of our study.

Standard life expectancy in Brazil has exhibited an upward trajectory, albeit with significant regional disparities. In Bahia, specifically, the analysis reveals a complex scenario, influenced by socioeconomic factors and public health determinants. Studies indicate that, despite advancements, challenges persist regarding premature mortality from chronic diseases and external causes, such as urban violence.^25^ Furthermore, life expectancy distribution varies considerably between urban and rural areas of the state, reflecting inequalities in access to healthcare services and basic sanitation.^26^ The GBD 2019 study corroborates these findings, highlighting the substantial burden of non-communicable diseases and interpersonal violence on YLL in Bahia, underscoring the necessity for targeted public health interventions.^27^ Our analysis, based on the GDB, demonstrates a perceptible trend of declining life expectancy with advancing age in both sexes. Notably, women consistently exhibit a higher life expectancy compared to men within each age cohort. This disparity underscores the influence of biological and sociocultural determinants on mortality patterns. The progressive reduction in life expectancy across all age groups suggests an increased vulnerability to mortality as individuals age, highlighting the importance of age-specific public health interventions.

The DM significantly contributes to premature mortality, with its impact extending beyond direct mortality to encompass substantial YLL.^28^ This condition accelerates the onset of comorbidities, such as cardiovascular diseases and renal failure, thereby reducing life expectancy.^29^ The burden of premature mortality attributable to DM varies across populations, influenced by socioeconomic factors and access to healthcare.^30^ In our study, the temporal analysis of YLL attributable to premature mortality due to diabetes mellitus in Bahia, Brazil, revealed an upward trend over the study period. This escalation suggests a progressive increase in the disease’s impact on the population’s life expectancy. The observed fluctuations indicate potential influences from evolving healthcare access, socioeconomic changes, and public health interventions, particularly during the COVID-19 pandemic period.

The gender disparities in YLL due to premature mortality from DM highlight significant variations in the disease’s impact. Studies consistently report a higher prevalence of YLL among men, potentially attributable to differential risk factors and neglect in accessing healthcare.^31^ Biological factors, such as hormonal differences and genetic predispositions, may also contribute to these disparities.^32^ Additionally, sociocultural determinants, including lifestyle behaviors and occupational exposures, influence disease progression and mortality.^33^ Addressing these gender-specific factors is of paramount importance for developing targeted interventions aimed at reducing premature mortality due to DM. Our analysis of age-standardized premature mortality rates due to DM in Bahia revealed a persistent gender disparity. In our study, men exhibited a higher prevalence of mortality compared to women, consistent with data reported in the literature. While both genders displayed fluctuations, men demonstrated a more pronounced upward trend, particularly after 2014. This suggests a greater vulnerability among men to DM-related complications, potentially influenced by neglect in seeking medical care or lifestyle-related factors.

Premature mortality due to DM carries significant socioeconomic implications, disproportionately affecting individuals in lower socioeconomic strata. Studies indicate that DM-related premature deaths contribute to substantial productivity losses, as they predominantly occur during working-age years, exacerbating economic burdens on families and healthcare systems.^34^ Additionally, individuals from disadvantaged backgrounds often face barriers to accessing timely and effective healthcare, leading to delayed diagnoses and suboptimal disease management.^35^ This perpetuates a cycle of health inequities, where socioeconomic determinants such as education, income, and occupation directly influence disease outcomes.^36^ Addressing these disparities requires targeted public health interventions, including improved healthcare access, education on preventive measures, and policies aimed at reducing socioeconomic inequities, to attenuate the broader societal impact of premature DM mortality. Based on the results of our study, it is evident that financial loss, coupled with declining per capita income, underscores economic strain. The disproportionate impact on the working-age population, particularly exacerbated during the 2020 pandemic, highlights the fragility of chronic disease management during health crises. This results in productivity loss, family instability, and increased reliance on social support, exacerbating pre-existing economic disparities in the region. Thus, premature mortality due to DM in Bahia, as evidenced by the rise in YLL, imposes substantial socioeconomic burdens.

Thus, our analysis of YLL due to premature mortality from DM in Bahia, Brazil, underscores a public health challenge. The rising YLL rates, coupled with pronounced gender disparities and socioeconomic impacts, demand immediate and targeted interventions. By acknowledging the complex interplay of demographic, biological, and sociocultural factors, public health services can devise strategies to attenuate premature mortality among diabetic individuals, thereby improving population health and reducing economic and social costs.

## CONCLUSION

Analysis of Bahia’s demographic data, utilizing IBGE population estimates and GBD 2019 metrics, reveals an escalating trend in YLL due to premature DM mortality. This increase, particularly pronounced post-2015 and exacerbated by the 2020 pandemic, highlights a significant public health challenge. Gender disparities, with males exhibiting higher mortality, and substantial socioeconomic impacts, including lost productivity and economic strain, necessitate integrated public health and economic interventions to attenuate this burden.

## Data Availability

All data produced in the present work are contained in the manuscript

